# Transmission dynamics of the COVID-19 epidemic in India and modelling optimal lockdown exit strategies

**DOI:** 10.1101/2020.05.13.20096826

**Authors:** Mohak Gupta, Saptarshi Soham Mohanta, Aditi Rao, Giridara Gopal Parameswaran, Mudit Agarwal, Mehak Arora, Archisman Mazumder, Ayush Lohiya, Priyamadhaba Behera, Agam Bansal, Rohit Kumar, Ved Prakash Meena, Pawan Tiwari, Anant Mohan, Sushma Bhatnagar

**Affiliations:** All India Institute of Medical Sciences (AIIMS), New Delhi; Indian Institute of Science Education and Research (IISER), Pune; Super Specialty Cancer Institute & Hospital, Lucknow; All India Institute of Medical Sciences (AIIMS), Bhubaneswar; Cleveland Clinic, Ohio, USA

## Abstract

**Background:** The SARS-CoV-2 pandemic has quickly become an unprecedented global health threat. India with its unique challenges in fighting this pandemic, imposed one of the world’s strictest and largest population-wide lockdown on 25 March 2020. Here, we estimated key epidemiological parameters and evaluated the effect of control measures on the COVID-19 epidemic in India. Through a modelling approach, we explored various strategies to exit the lockdown.

**Methods:** We obtained data from 140 confirmed COVID-19 patients at a tertiary care hospital in India to estimate the delay from symptom onset to confirmation and the proportion of cases without symptoms. We estimated the basic reproduction number (R_0_) and time-varying effective reproduction number (R_t_) after adjusting for imported cases and reporting lag, using incidence data from 4 March to 25 April 2020 for India. We built upon the SEIR model to account for underreporting, reporting delays, and varying asymptomatic proportion and infectivity. Using this model, we simulated lockdown relaxation under various scenarios to evaluate its effect on the second wave, and also modelled increased detection through testing. We hypothesised that increased testing after lockdown relaxation will decrease the epidemic growth enough to allow for greater resumption of normal social mixing thus minimising the social and economic fallout.

**Results:** The median delay from symptom onset to confirmation (reporting lag) was estimated to be 2·68 days (95%CI 2·00–3·00) with an IQR of 2·03 days (95%CI 1·00–3·00). 60·7% of confirmed COVID-19 cases (n=140) were found to be asymptomatic. The R_0_ for India was estimated to be 2·083 (95%CI 2·044–2·122; R^2^ = 0·972), while the R_t_ gradually down trended from 1·665 (95%CI 1·539–1·789) on 30 March to 1·159 (95%CI 1·128–1·189) on 22 April. In the modelling, we observed that the time lag from date of lockdown relaxation to start of second wave increases as lockdown is extended farther after the first wave peak. This benefit was greater for a gradual relaxation as compared to a sudden lifting of lockdown. We found that increased detection through testing decreases the number of total infections and symptomatic cases, and the benefit of detecting each extra case was higher when prevailing transmission rates were higher (as when restrictions are relaxed). Lower levels of social restrictions when coupled with increased testing, could achieve similar outcomes as an aggressive social distancing regime where testing was not increased.

**Conclusions:** The aggressive control measures in India since 25 March have produced measurable reductions in transmission, although suppression needs to be maintained to achieve sub-threshold R_t_. Additional benefits for mitigating the second wave can be achieved if lockdown can be feasibly extended farther after the peak of active cases has passed. Aggressive measures like lockdowns may inherently be enough to suppress the epidemic, however other measures need to be scaled up as lockdowns are relaxed. Expanded testing is expected to play a pivotal role in the lockdown exit strategy and will determine the degree of return to ‘normalcy’ that will be possible. Increased testing coverage will also ensure rapid feedback from surveillance systems regarding any resurgence in cases, so that geo-temporally targeted measures can be instituted at the earliest. Considering that asymptomatics play an undeniable role in transmission of COVID-19, it may be prudent to reduce the dependence on presence of symptoms for implementing control strategies, behavioral changes and testing.

## Introduction

Originating out of Wuhan, China in December 2019,^1^ the coronavirus disease 2019 (COVID-19) was declared a pandemic by the WHO on March 11 2020.^2^ As of 2 May 2020, there have been more than 3 200 000 cases and 230 000 deaths worldwide, and close to 40 000 cases and 1200 deaths in India.^3^ India reported its first COVID-19 case on 30 January 2020, although actual epidemic growth started from early March.^4^

For any novel infectious disease, the scale of its public health impact is determined by the basic reproduction number ‘R_0_’ which is the average number of secondary infections generated by an infectious index case in a wholly susceptible population. The R_0_ of an infection determines its potential to start an outbreak, the severity of control measures needed to contain the spread, and the fraction of the population that will be infected in the absence of interventions.^5^ However, once an outbreak is underway, the time-varying effective reproduction number ‘R_t_’ is more relevant as it tracks the subsequent changes in transmission, and can thus be used to monitor the efficacy of control measures and adjust them accordingly.^6–8^ However, any given transmission event is reflected in the data only after a delay, which must be accounted for in the estimation of such indicators for accurate interpretation.^7^ Previous studies have shown that a severe epidemic with R_0_~2·4 can be contained by combining effective quarantine, behavioral change to reduce social mixing, targeted antiviral prophylaxis, and pre-vaccination.^5^ However, in the absence of targeted therapeutics and vaccination for COVID-19, an unprecedented one-third of the world’s population is currently under lockdowns – with the primary target of reducing the R_t_ below the threshold of 1.^6,9^

India responded to the COVID-19 pandemic rapidly and decisively by imposing a nation-wide lockdown on 25 March 2020, when there were 536 cases and 10 deaths.^3,10^ This ‘suppression strategy’, though effective, has its limitations- the social and economic cost of such population-wide social distancing is huge, which limits the long-term implementation of these measures.^9,11^ Additionally, containing COVID-19 in India is a unique challenge due to its high population density, underprepared healthcare system, and wide socio-economic disparity.^12^ A large proportion of India’s labour force works as daily-wage laborers or migrant workers, and are especially affected during such times, making lockdowns untenable without parallel social support.^13^ There could be yet unseen adverse effects in the form of non-COVID-19 morbidity and mortality due to aggravation of malnutrition, chronic diseases and lack of access to healthcare during this time.^14^ At the same time, premature withdrawal of lockdowns without adequately planned interventions for the post-lockdown phase may lead to re-emergence later, or the second wave. ^6,9^ Thus there arises a need to create a balance to ensure that the disease is contained and the healthcare system remains well prepared while minimizing the collateral damage from intensive blanket interventions. Comprehensive lockdown exit strategies will be central to the future course of the pandemic.^10^ In such scenarios with limited primary information, dynamic mathematical models can provide actionable insights for researchers and policymakers.^7,9,15^

Evidence suggests that COVID-19 has a wide clinical spectrum which ranges from asymptomatic to fatal infections, which coupled with high infectivity can lead to a large number of infections and deaths.^16^ It may be possible that COVID-19 transmission is driven significantly by undetected asymptomatics while fatality is driven by severe cases- a devastating combination.^17,18^ Some have deemed asymptomatic transmission as the “Achilles’ Heel” of the current control strategies against COVID-19,^19^ and it is important to consider the range of uncertainty regarding same when simulating COVID-19 transmission.

In this study, we estimate the key transmission parameters for COVID-19 in India and its states, and analyse how interventions affected transmission levels across time. Considering that blanket lockdowns are an initial rather than a final step in controlling this pandemic, we model the effect of relaxing public health interventions at various time-points. We evaluate the impact of increased detection of infections in the community through expanded testing strategies in containing transmission when restrictions are relaxed.

## Methods

### Data sources

For estimating the proportion of asymptomatic cases and the delay from symptom onset to confirmation, we obtained data from 140 COVID-19 patients admitted to a tertiary care hospital near Delhi, India (appendix p5). For estimating the basic reproduction number (R_0_) and effective reproduction number (R_t_) for India and various states, we used data from COVID19India from 04 March to 25 April 2020, which is curated based on multiple verified sources.^4^ For model fitting and parameter estimation, we used time-series data for India from Johns Hopkins University COVID-19 database, from 16 March to 18 April 2020.^3^ A laboratory confirmed case irrespective of symptoms is counted as a confirmed COVID-19 case in India. Testing criteria are provided in appendix p9. We used the World Bank Population Database for population data for India.^20^

### Estimation of basic reproduction number (R_0_)

The best-fit R_0_ was calculated for the national and state level incidence data using the *R_0_ package* in R 3·6·3 using two independent methods: Maximum Likelihood (ML) method and the Exponential Growth (EG) method after adjusting the incidence data for imported cases.^21–23^ We assumed the serial interval to be gamma distributed with a mean of 3·96 days (95% CI 3·53–4·39) and a SD of 4·75 days (95% CI 4·46–5·07), based on a large study of 468 infector-infectee pairs in China.^24^ We analysed the sensitivity of the estimated R_0_ to the choice of the time period over which the R0 was estimated and the serial interval (appendix p8).

### Estimation of reporting lag, lag adjusted incidence, and time-varying effective reproduction number (R_t_)

A variable delay occurs from symptom onset to case confirmation (henceforth referred to as the reporting lag) which is attributed to multiple factors including time taken to seek care (patient dependent) and time taken to detect and test the case (healthcare-system dependent). Since all included patients were tested and confirmed positive within a day of hospitalization, the time from symptom onset to hospitalization obtained from the data approximates the reporting lag of these cases. We assume these to be same for the purpose of our study. Due to lack of data, we assume that the reporting lag for India and each state is statistically the same as the estimated reporting lag for the 53 patients from Delhi whose onset date was known. For each reported case, onset dates were sampled to generate 1000 lag-adjusted datasets for incidence by onset (appendix p5-6) from which, the time-varying R_t_ was calculated using *EpiEstim* package in R 3·6·3 which uses the Time Dependent Maximum Likelihood approach.^25,26^ The same serial interval distribution was used as for R_0_ estimation.^24^ We determined both the import adjusted *R_t_* and unadjusted *R_t_* for India, where cases in the national incidence data not explicitly labeled as ‘imported’ were considered to be locally transmitted. The *R_t_* trends were overlaid with major epidemic events and mobility data to analyse possible temporal correlations.^27^

### Modelling the pandemic using dynamic compartmental models

In order to model the spread of SARS-CoV-2 in the population, we generalize the extensively used SEIR model for infectious diseases,^28^ to account for (1) time lag from symptom onset to case being reported in data, (2) underreporting of actual infections due to testing constraints, (3) varying proportion of infections being asymptomatic, (4) varying infectivity levels of asymptomatics, and (5) time dependent effect of implementing and relaxing control measures. For introducing the required complexities, we build a model as shown in Figure 1. Model parameters are defined in Table 1. Through a positive protection rate (α), the susceptible population gradually decreases to account for the effect of increasingly intensive social distancing policies and improved public behaviour in reaction to the epidemic.^36^ We introduce a deprotection rate (σ) which increases the susceptible pool once social distancing policies are relaxed. We set the probability of infected case being asymptomatic (p_a_) to 0·2, 0·4, 0·6, 0·8, as reported estimates for the percent of infections that are asymptomatic range widely from 18%-80%.^17,18,32–34^ We consider that asymptomatic cases do not exhibit coughing, sneezing or sputum production and are thus expected to show lower infectivity than symptomatics. We conservatively assume an asymptomatic patient is 50% as infective as a symptomatic patient (a_i_ =0·5). We set the fraction of detected asymptomatics (f_a_) at baseline to 0·1,^35^ the incubation period (*γ^−^*^1^) to 5·1 days,^29^ and the infectious period for asymptomatics (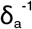) to 8 days based on virologic and epidemiologic studies.^30,31^ We assume that all symptomatics are detected, and that no asymptomatic dies from the disease. Further details of the model, including the governing equations are available in appendix p3-4.

**Table 1:** Parameters for SEIR-QDPA model.

| PARAMETER | VALUE | REFERENCE |
| --- | --- | --- |
| Protection rate ($\alpha$) | - | Estimated |
| Deprotection rate ($\sigma$) | **0.66** for fast lockdown relaxation (largest possible value for stable simulation) | Tested (Fig 4) |
| Transmission rate ($\beta$) | - | Estimated |
| | **0.9, 0.8, 0.7, 0.6, 0.5, 0.3** of estimated $\beta$ for social mixing levels | Tested (Fig 5) |
| Median incubation period ($\gamma^{-1}$) | **5.1** days | Lauer et al <sup>29</sup> |
| Reporting lag for symptomatic ($\delta_s^{-1}$) | - | Estimated |
| Infectious period for asymptomatic ($\delta_a^{-1}$) | **8** days | Wolfel et al, He et al <sup>30,31</sup> |
| Mortality rate ($\kappa$) | - | Estimated |
| Recovery rate ($\lambda$) | - | Estimated |
| Infectivity of asymptomatic compared to symptomatic ($a$) | **0.5** (0.25, 0.75 for sensitivity analysis) | Assumed |
| Probability of infected case being asymptomatic ($p_a$) | **0.2, 0.4, 0.6, 0.8** | This study, Mizumoto et al, Gudbjartsson et al, Lavezzo et al, Media reports <sup>17,18,32-34</sup> |
| Probability of detection of asymptomatic case ($f_a$) | **0.1** (0.05, 0.2 for sensitivity analysis) | Russel et al <sup>35</sup> |
|  | **0.2, 0.3, 0.4, 0.5, 0.6, 0.8** for increased testing | Tested (Fig 5) |

Sensitivity analysis to the choice of assumed parameters $a$ , $f_a$ , and $p_a$ was performed for the fitted parameters $\alpha$ , $\beta$ , $\delta_s^{-1}$ , $\kappa$ and $\lambda$ in appendix p16-18. Confidence intervals for incubation period and asymptomatic infectious period have not been included since these parameters are fixed and not sampled.

**Figure 1:**
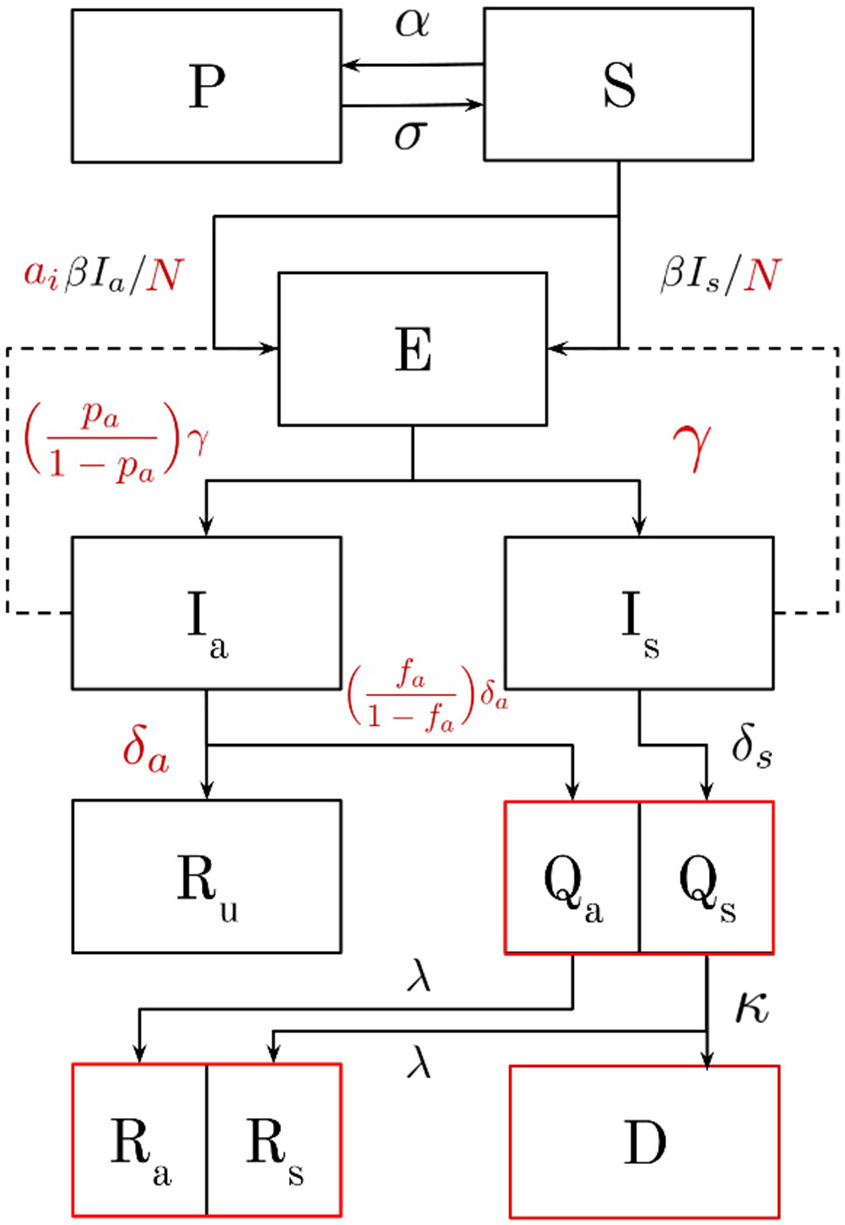
Schematic for SEIR-QDPA model. Compartments include S(susceptible), E(infected but not yet infectious), Ia(undetected asymptomatic; infectious), Is(undetected symptomatic; infectious), Qa(detected and quarantined asymptomatic), Qs(detected and quarantined symptomatic), Ru(undetected recovered asymptomatic), Ra(recovered detected asymptomatic), Rs(recovered detected symptomatic), D(dead), and P(protected; non-susceptible). Compartments in red are fitted to data; Q=Qa+Qs to active cases, R=Ra+Rs to cumulative recovered cases, and D to cumulative deaths. Transition rates in red are inputs to the model, while others are estimated (Table 1).

We estimated the unknown parameters of the model by fitting time-series data for active cases (= cumulative confirmed cases – cumulative recoveries – cumulative deaths), cumulative recoveries and cumulative deaths to the Q(t) = Q_s_(t) + Q_a_(t), R(t) = R_s_(t) + R_a_(t), and D(t) compartments respectively. We fitted for the values of transmission rate (β), protection rate (α), reporting lag (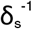), recovery rate (λ), mortality rate (κ), and initial exposed and infected individuals (E_0_ and I_0_). As a counterfactual, we explored the size and temporality of the first wave in an ideal scenario where strict control measures could be maintained for long periods, by simulating the model with the estimated parameters. This continually enforces the estimated protection rate thus assuming that control measures continue with initial stringency till the end of simulation. Here, we defined three key time points which are inherent to epidemic progression-time at peak of daily new reported cases (t1), time at peak of active cases (t2), and time when recovered cases > active cases (t3). Sensitivity of our results to assumptions of p_a_, f_a_ and a_i_ was analysed.

The following terms in the article signify values obtained by combining multiple compartments: ‘Symptomatic cases’- cumulative symptomatic cases once detected; ‘Detected cases’- cumulative detected cases including symptomatic and asymptomatic cases; ‘Total infections’- cumulative infections including detected cases and undetected asymptomatic infections.

### Simulating the effect of lockdown relaxation

To model complete lifting of the nation-wide lockdown, α was set to zero and σ was set to a large value such that the entire protected population was emptied into the susceptible population in a short interval (t_½_~1·0 day). We triggered this change on 4 May 2020 (tentative date of lockdown relaxation in India at time of study) and 7-day intervals thereafter, to compare outcomes if lockdown is lifted at different dates. We also considered the scenario where the lockdown is re-enforced after a fixed relaxation period which we modelled by setting σ to zero and re-enforcing the same α as before lockdown relaxation.

### Simulating the effect of increased testing

We hypothesised that increased testing after lockdown relaxation will decrease the epidemic growth enough to allow for greater resumption of normal social mixing, thus minimising the social and economic fallout resulting from vigorous restrictions. To model the effect of increased testing capacity and subsequent improved detection, we assumed that it increased the asymptomatic detection rate f_a_ from 0·1 in lockdown, to 0·2, 0·3, 0·4, 0·5, 0·6, and 0·8 after lockdown relaxation starting May 4. An alternate interpretation of testing which is independent of clinical severity is discussed in appendix p4. To model the effect of varying levels of reduced social mixing and positive behavior change after the lockdown is lifted, we changed the transmission rate to 90%, 80%, 70%, 60%, 50% and 30% of the original β, starting May 4. Varying levels of social mixing mainly influences the contact rate, while behaviour changes like wearing masks and hand washing decrease transmissibility given a contact- the transmission rate β captures both these changes.

## Results

### Basic Reproduction Number ‘R_0_

The exponential growth (EG) method had a better best-fit R^2^ over a larger time period and was less sensitive to the choice of the time period (appendix p7-8). The best fit R_0_ for India was found to be 2·083 (95% CI 2·044–2·122; R^2^ = 0·972). Taking into consideration the uncertainty in reported serial intervals (SI), the R_0_ ranged from 2–2·5 for SI ranging from 4–4·6 days.^24,37–39^ Results were found to be sensitive to the SI distribution, thus we report R_0_ based on reliable SI estimates from 468 infector-infectee pairs in China,^24^ and also consider a range of possible SI’s based on other studies.^37–39^ The R_0_ estimates for various states of India have been provided in appendix p7.

### Reporting lag, lag adjusted incidence, and time-varying effective reproduction number ‘R_t_’

Out of a 140 laboratory confirmed COVID-19 patients, 85 (60.7%) were asymptomatic while 55 (39.3%) were symptomatic. For 53 symptomatic patients, the reporting lag was found to have a mean of 3·40 days (95% CI 2·87–3·96) with SD of 2·09 days (95% CI 1·52–2·56) and a median of 2·68 days (95% CI 2·00–3·00) with IQR of 2.03 days (95% CI 1.00–3.00). The gamma distribution with shape parameter 3·45 (95% CI 2·42–5·19) and rate parameter 1·02 (95% CI 0·70–1·60) was the best fit to the distribution (appendix p5-6).

The first cases of local transmission in India were reported on 4 March, who were family members of an initial imported case. The number of imported cases in India started increasing from early-March, peaked a day after the international travel ban on 23 March,^40^ and gradually came to a halt on 5 April with a total of 546 imported cases (Figure 2A). Incidence by onset and time-varying R_t_ could be ascertained upto 22 April 2020 since some cases with onset after this date may not have been reported yet in the data, due to the reporting lag. The R_t_ trends for India showed visible fluctuations over time (Figure 2B). The first uptick in unadjusted R_t_ (blue band) starting around 13 March 2020 was presumed to be an artifact due to imported cases, since it coincided with increasing imported case onsets, and was not accompanied by a concurrent uptick in import adjusted R_t_ (pink band). The second uptick in unadjusted R_t_ correlated with the rise in adjusted R_t_, indicating that local transmission was driving this rise. This rise started around the imposition of the nationwide lockdown on 25 March and peaked on 30 March at an adjusted R_t_ of 1·665 (95%CI 1·539–1·789). After this peak, the R_t_ continued to decrease to 1·300 (1·247–1·353) on 8 April, 1·213 (1·175–1·251) on 15 April and further to its lowest yet value of 1·159 (1·128–1·189) on 22 April. A sharp dip in mobility is noted at the voluntary public curfew on 22 March, which is sustained after the nationwide lockdown was enforced on 25 March, except for a rise in residential neighbourhood mobility (Figure 2C). The daily R_t_ values for India, and R_t_ trends for states of India are provided in appendix p11-14.

**Figure 2:**
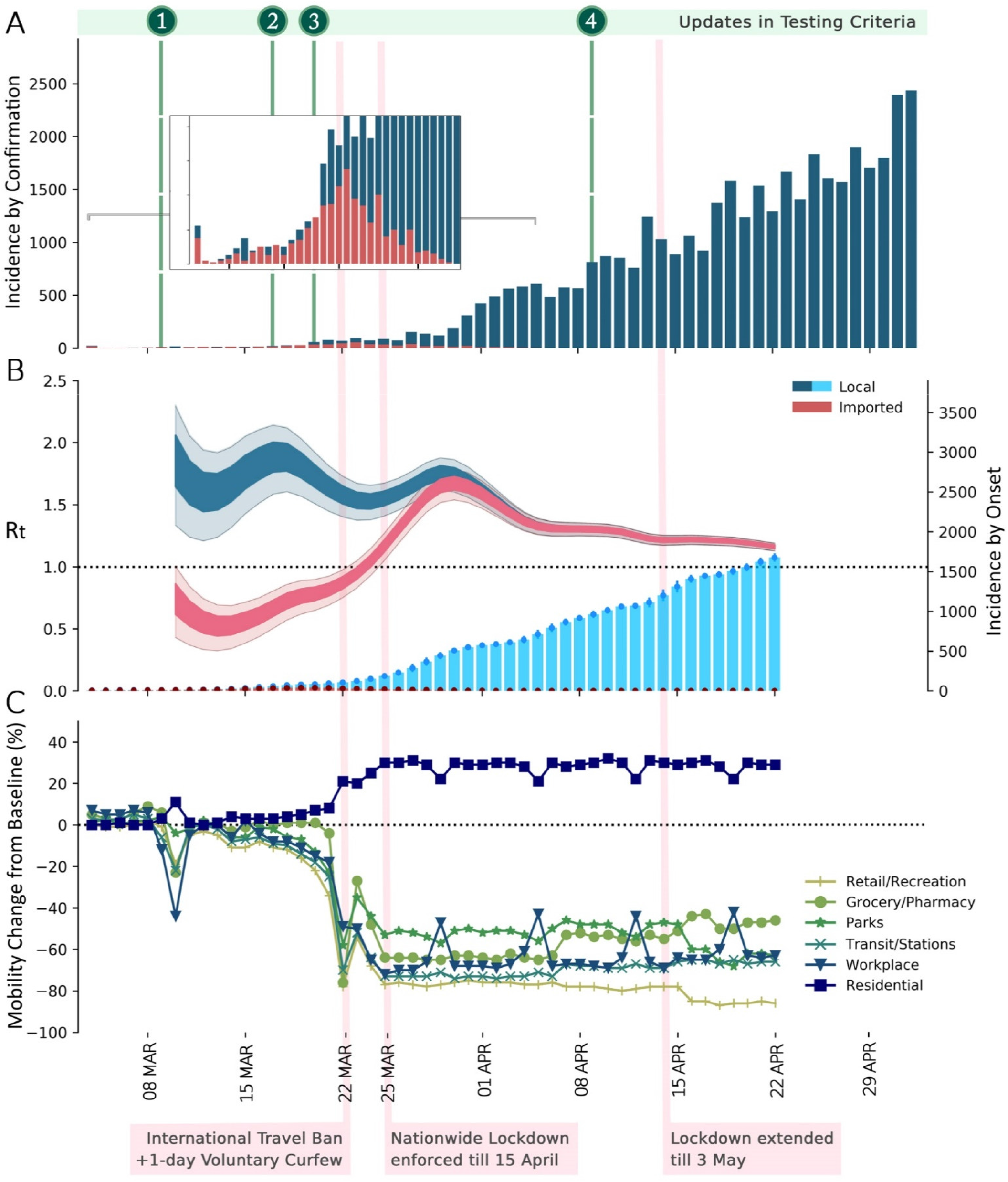
Transmission dynamics and incidence of COVID-19 in India, overlaid with major events and mobility trends. **[A]** Daily new cases by confirmation date in India up to 2 May 2020 stratified as imported (red) and local (dark blue). The dates of testing criteria updates are overlaid since the scope of testing influences the number of confirmed cases (appendix p9).^41^ **[B]** Daily new cases by onset date (estimated epidemic curve) up to 22 April 2020 in India stratified as imported (red) and local (light blue); and the time-varying effective reproduction number R_t_ adjusted for importations (pink) and without adjusting for importations (blue), over 5-day windows. Dark bands indicate 50% CI and light bands indicate 95% CI for estimated R_t_. Similar graphs for states of India are provided in appendix. **[C]** Mobility trends in India, compared to a baseline median value for the corresponding day of the week, during the 5-week period Jan 3–Feb 6, 2020. Holiday due to the Holi festival on 10 March 2020 caused a dip in mobility. A sharp dip in mobility is noted at the voluntary public curfew on 22 March and after the nationwide lockdown was enforced on 25 March, except for a rise in residential neighbourhood mobility. The weekly rise in workplace mobility appears to be an artifact due to comparison with normal weekends at the baseline. Source-Google community mobility reports.^27^ Major interventions are shown, the effects of which are best correlated with R, trend and mobility changes, since these changes occur in real-time. R_t_= time-varying effective reproduction number

### Estimated model parameters and ideal first wave scenario

The model was able to fit the data well for the early exponential phase of the growth and also captured the recent slowdown in epidemic growth through the protection rate. The estimated model parameters for the range of assumptions are provided in appendix p15-18. The reporting lag estimated from model fit ranged from 2·08 to 2·53 days, which is similar to the median reporting lag of 2·68 days estimated from primary patient data. Assuming 60% of total infections to be asymptomatic (p_a_ = 0·6) and that control measures continued with initial stringency, our model predicted the first wave (Figure 3) at 19741 (95% CI 17187–21870) maximum active cases, 34049 (30330–38648) cumulative detected cases of which 29610 (26377–33609) were symptomatic, till the end of first wave. The total infections were 73820 (65746–83811). Key time points were predicted as - time at peak of daily new reported cases (t1) between 16 April and 19 April 2020, time at peak of active cases (t2) between 3 May and 8 May 2020, and time when recovered cases > active cases (t3) between 25 May and 29 May 2020. 95% CI of 1000 bootstrapped predictions are reported here. The epidemic size increased and key points were delayed with higher assumptions of asymptomatic proportion (appendix p19). It is important to note that the model does not consider a parallel leakage of protected compartment back into the susceptible compartment, which tends to happen in reality as lockdowns are not perfect. In addition, control measures can not be practically maintained indefinitely with initial stringency. Thus, these first wave estimates will obviously be lower than actual data in future, however, they give us the valuable opportunity to model various interventions and explore alternate scenarios.

**Figure 3:**
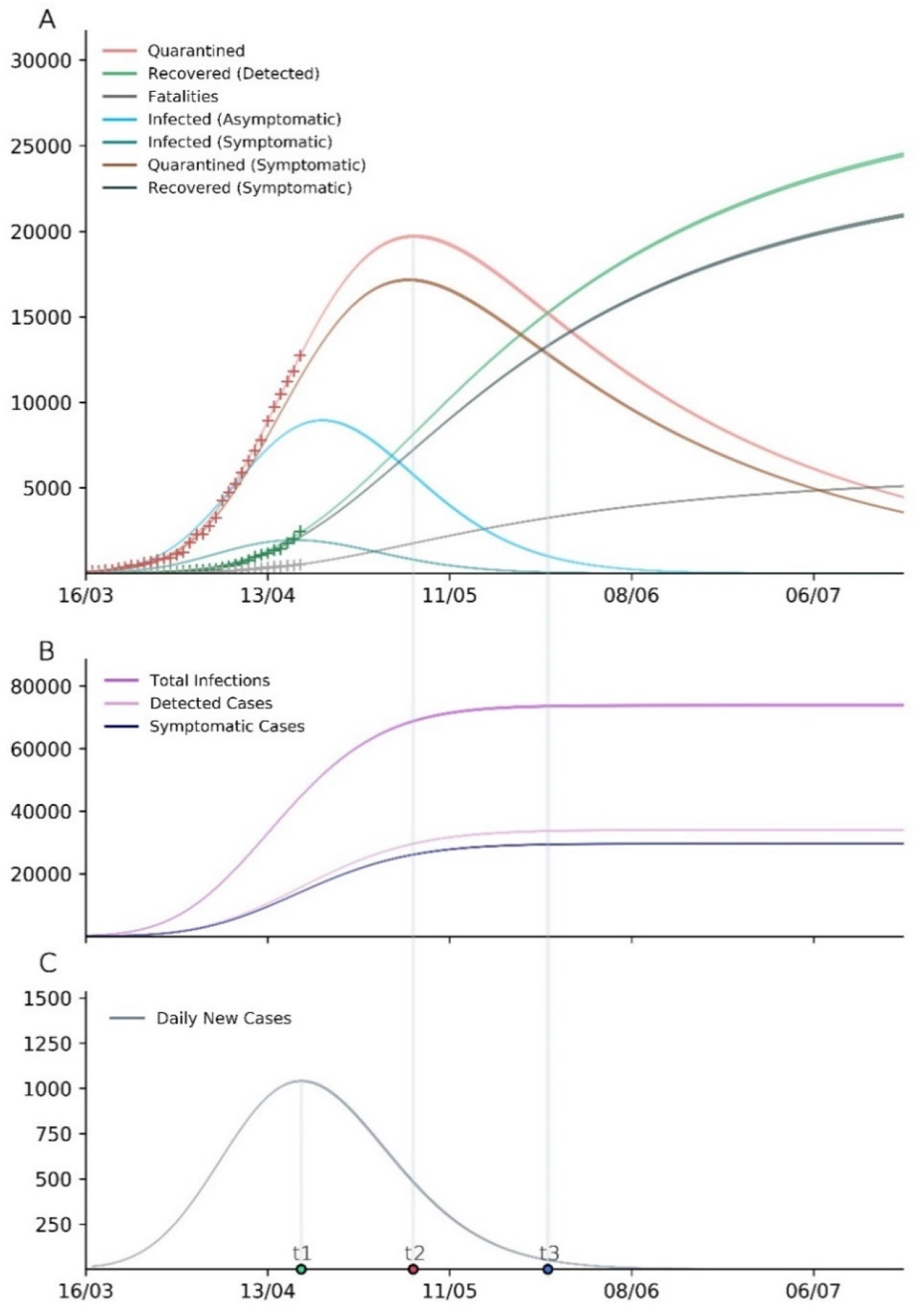
Model simulation of the first wave of COVID-19 in India assuming the lockdown continues indefinitely with the initial stringency. **[A]** Simulated values of model compartments over time. Quarantined cases are equivalent to the active cases at a particular time. ‘+’ represents data with which the model was trained. **[B]** Predicted total infections, detected cases, and symptomatic cases over time. **[C]** Predicted daily new cases over time. Bands represent 95% CI for the mean prediction over 1000 bootstraps. Three key time points in epidemic progression are shown-time at peak of daily new reported cases (t1), time at peak of active cases (t2), and time when recovered cases > active cases (t3). Results shown for the baseline assumptions (asymptomatics are 50% infectious compared to symptomatics, 60% of total infections are asymptomatic, 10% asymptomatics are detected and quarantined). Results for other assumptions in appendix.

### Impact of lockdown relaxation and its temporality

On a complete removal of the lockdown irrespective of the date of relaxation, we observed that the number of active cases will start to rise exponentially after a variable delay (Figure 4A). We observed that delaying the lockdown relaxation increases the time lag from the date of relaxation to the date of new rise in active cases (start of second wave), in a linear fashion with Pearson’s R = 0·997 (95% CI 0·989–0·999; p<0·0001) as shown in Figure 4B. When we simulated limited time duration relaxation periods, we found that there is a rise in active cases observed in all scenarios but the extent of the rise is highly dependent on when the relaxation was started and the duration of the relaxation period (Figure 4C and 4D). Both delays in the lockdown relaxation and shorter relaxation periods reduced the number of active cases at the peak. In the case of a gradual lockdown release, the second wave was smaller and further delayed when compared to sudden relaxation (appendix p26-27). The result shown in Figure 4 considers that the peak of active cases is on 4 May 2020, while the actual peak of active cases may occur later than 4 May, as discussed earlier. These findings may be generalized to any first wave scenario, when interpreted with respect to the actual peak date (which will be comparable to red point in Figure 4B), instead of the absolute dates that are simulated.

**Figure 4:**
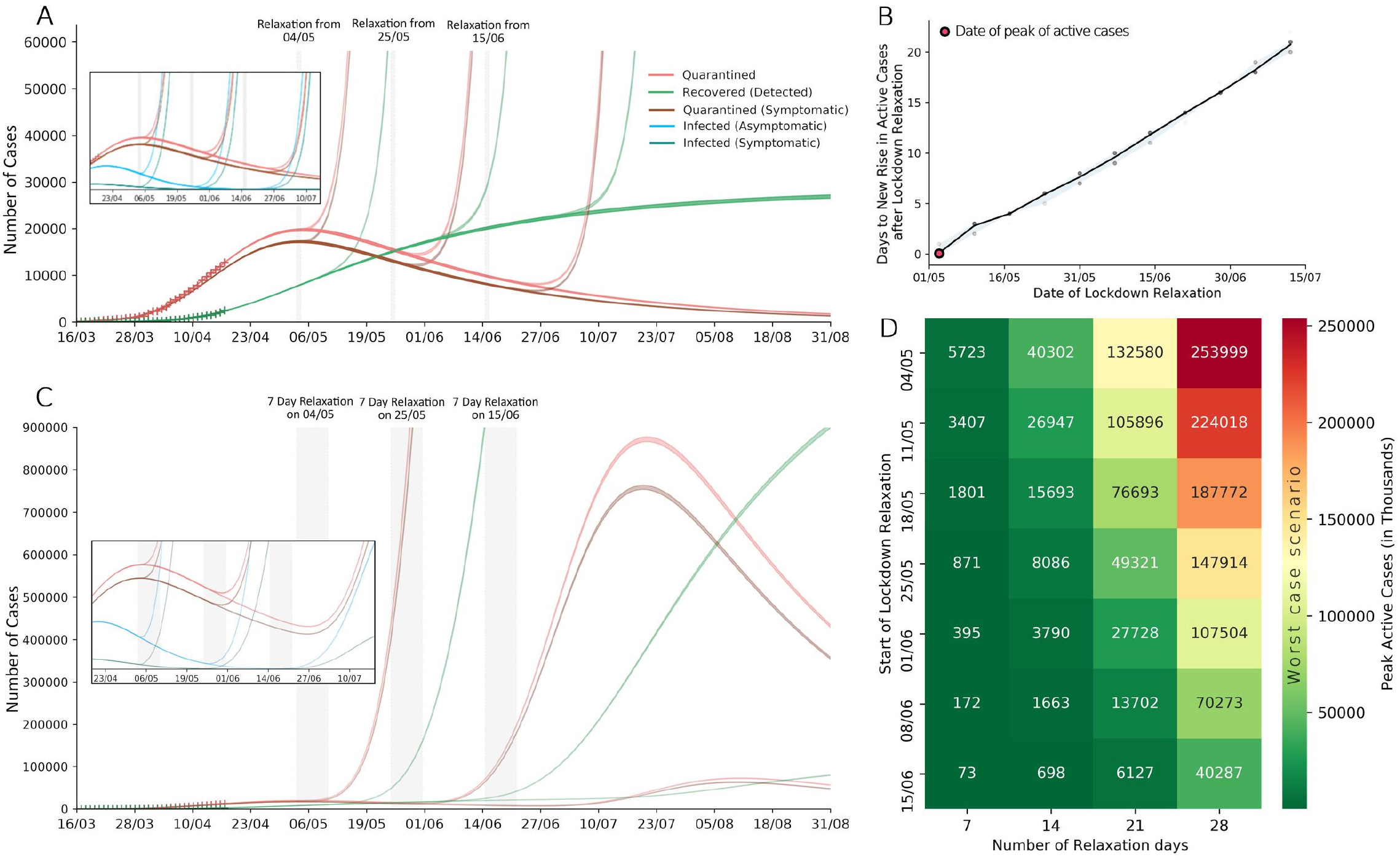
Effect of complete lockdown relaxation under various scenarios. Results are shown for the baseline assumptions (asymptomatics are 50% infectious compared to symptomatics, 60% of total infections are asymptomatic, 10% of asymptomatics are detected and quarantined). Bands represent 95% CI for the mean prediction over 1000 bootstraps for all model plots. **[A]** Simulated values of the model compartments Q (active cases), Q_s_ (active symptomatic cases) and R (recovered) under complete and sustained lockdown relaxation starting 4 May, 25 May, and 15 June 2020; showing increasing delay to start of the second wave with later relaxation. Inlay shows the underlying depletion of undetected infectious pool as the first wave crosses the peak. **[B]** Days to new rise in active cases (time delay after respective relaxation date) at different dates for lockdown relaxation. This effect is expected to be generalizable when interpreted with respect to the actual date of peak of active cases (compare with red point). Black line represents the line joining the mean lag for 1000 bootstrapped simulations, and bands represent 95% CI. **[C]** Simulated values of the model compartments Q (active cases), Q_s_ (active symptomatic cases) and R (recovered) under complete relaxation lasting 7 days, starting 4 May, 25 May, and 15 June 2020; showing increasing delay to start of the second wave and lower magnitude of the second wave with later relaxation. **[D]** Heatmap for the peak active cases under different lockdown relaxation durations and dates of start of relaxation. These are hypothetical worst-case values, where lockdown has been completely lifted across the country at once.

### Effect of increased testing on epidemic size and restoration of normal social mixing

Increased detection through expanded testing resulted in a decrease in the number of total infections and symptomatic cases (Figure 5A and 5B). The number of detected cases may remain almost constant at various levels of testing, due to a concurrent decrease in total infections and increase in the fraction of infections that were detected (ascertainment rate). However, a lower proportion of detected cases are symptomatic at higher testing, which highlights the significance of detecting more asymptomatic-mild infections.

**Figure 5:**
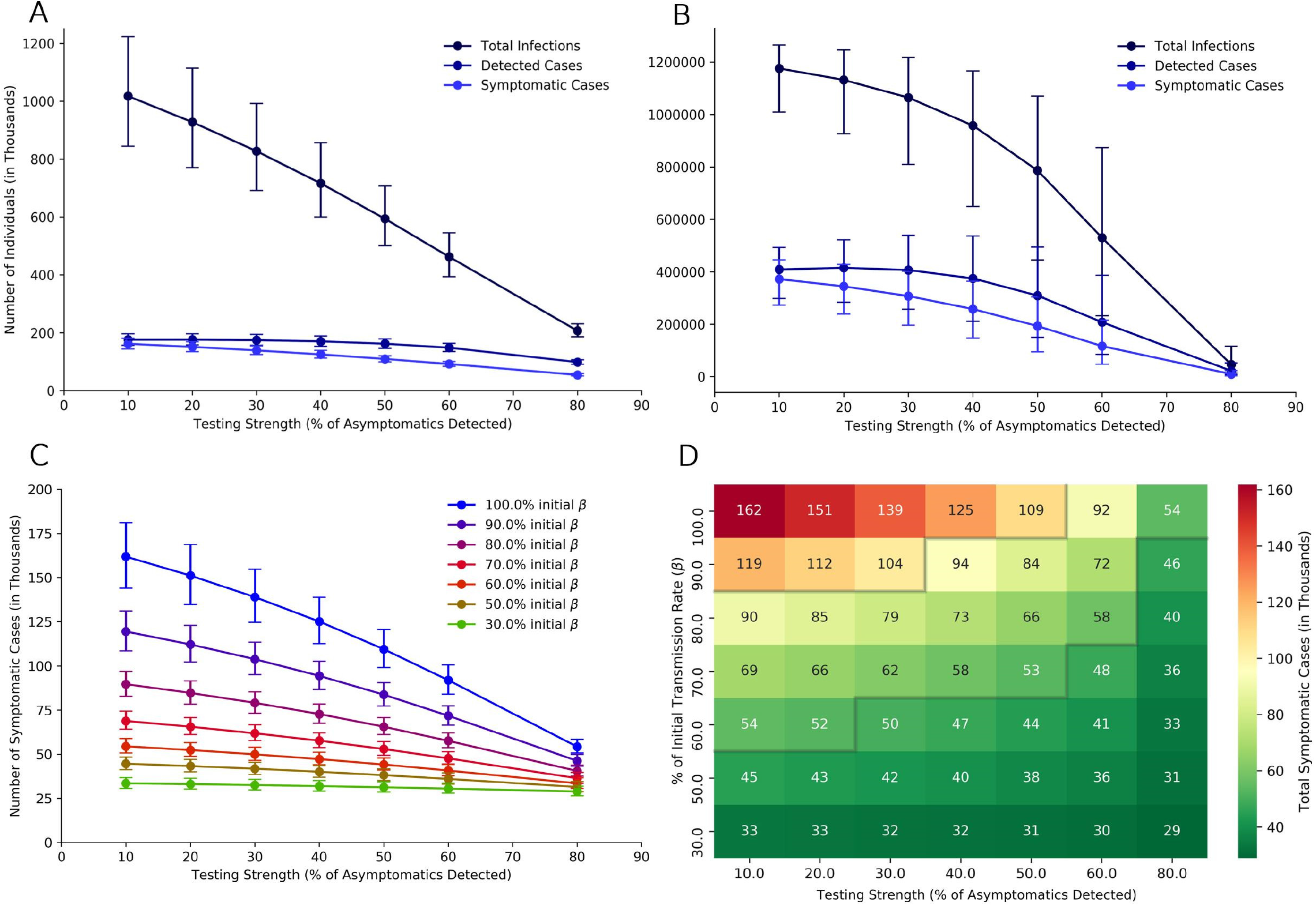
Effect of expanded testing and varying social mixing after complete lockdown relaxation. Results shown for the baseline assumptions (asymptomatics are 50% infectious compared to symptomatic, 60% of total infections are asymptomatic, 10% asymptomatics are detected before lockdown relaxation). Any increase in testing or any decrease in social mixing starts from the day of lockdown relaxation. Results for other assumptions in appendix. Error bars represent 95% CI for 1000 bootstrapped predictions. All values are given in thousands of individuals. **[A] and [B]** Total number of infections, detected cases and symptomatic cases at 15 days and 45 days after lockdown relaxation with varying levels of testing. **[C]** Effect of increasing testing (along x-axis) and decreasing social mixing (lines from top to bottom) on the number of symptomatic cases at 15 days after the lockdown relaxation. **[D]** Heatmap for total symptomatic cases after 15 days under different reductions in transmission rate (proxy for social distancing policies) and asymptomatic detection rate (proxy for testing policy). An example of a feasible combination of testing and social distancing policy is indicated by the area between two watershed lines (grey) for a containment target of 50,000-100,000 cases. Similar heatmap for total infections is given in appendix p24.

We further found that the positive impact of increased testing becomes more prominent at progressively higher values of transmission rate p (Figure 5C). At baseline assumptions (asymptomatics are 50% infectious compared to symptomatic, 60% of total infections are asymptomatic), when detection increased from 10% to 20% for 15 days– for every extra asymptomatic detected by increased testing, the number of infections prevented are 8·3 at β, 6·4 at 0·8 β and 3·5 at 0·5 β, while the number of symptomatic cases prevented are 1 at β, 0·9 at 0·8 β, 0·6 at 0·5 β. This benefit increases further with increases in testing above 20%. For an increase in detection from 10% to 20%, the symptomatic cases decreased by 6·6% at β, by 5·5% at 0·8β, and by 3% at 0·5β. For an increase in testing from 10% to 50%, the symptomatic cases decreased by 32·3% at β, by 26·8% at 0·8 β, and by 14·5% at 0·5β.

After lockdown relaxation, lower levels of social restrictions (high β) when coupled with increased testing, can achieve similar results as a more restrictive social distancing regime where testing was not increased (Figure 5D). That is, increased testing allowed greater resumption of normal social mixing after lockdown relaxation. An example of a feasible combination of testing and social restrictions is indicated by the area between two watershed lines (grey) in Figure 5D. Due to uncertainty in the percent of infections that are asymptomatic, we evaluated the effect of testing across the range of p_a_ (appendix p22-25).

## Discussion

The trend of effective reproduction number (R_t_) of COVID-19 in India indicates that control measures have been effective in slowing down the spread of COVID-19 across the country. To achieve sustained suppression, monitoring of the time varying R_t_ at district, state and national level should be done to reach and maintain an R_t_ close to the threshold value of 1. If lockdown is to be extended, additional benefits can be achieved if it is extended farther after the peak of active cases has passed. As these restrictions are relaxed, increased detection through testing will be essential in limiting the resurgence of cases and thus testing capacity should be ramped up preemptively before lifting restrictions. Considering that asymptomatics play an undeniable role in transmission of COVID-19, dependence on presence of symptoms for control strategies, behavioral changes and testing should be reduced.

The range of R_0_ of SARS-CoV-2 in India was found to be 2–2·5, with 2·083 being the best fit. Our results align with recent studies which estimate the R_0_ to be 2–2·7.^42–44^ In comparison, the R_0_ was 1·4–1·6 for the 2009 influenza (H1N1) pandemic, 2·0–3·3 for the 2003 SARS epidemic, and 2·0–3·0 for the 1918 Spanish flu pandemic, which reflects the seriousness of the current pandemic.^45-47^ The proportion of population that must become immune in order to halt the epidemic is given by 1 − 1/R_0_, the herd immunity threshold.^48^ For COVID-19, our estimates imply that approximately 50–60% of the population must be infected or vaccinated in order to attain long-term epidemic control. In reality, this threshold is usually higher due to non-homogenous mixing in populations.^48^

In early stages of the epidemic in India, we found that restrictions on international travel were effective in limiting the number of imported cases in India, although this is of limited importance once local chains of transmission had been established.^49^ Since testing of travelers was based on appearance of symptoms,^41^ asymptomatic imported infections that remained undetected may have played some role in the early spread of COVID-19.

A ‘suppression’ strategy (eg: lockdown) aims to arrest epidemic growth by reducing R_t_ below 1.^6,9^ After the nationwide lockdown was imposed on 25 March 2020, the mobility levels quickly dropped to low levels, but the R_t_ continued to increase till 30 March (Figure 2) probably due to inflation of estimated transmission by the Nizammudin cluster (a super-spreading event originating in Delhi)– which represented about 30% of total COVID-19 cases in India in early April, with latest data linking the cluster to 4291 cases across more than 15 Indian states.^50^ This event adds to the list of multiple COVID-19 super-spreader events around the world, which have caused unexpected spikes in cases.^51^ It should be noted that clusters may disproportionately inflate transmission estimates, because targeted testing of people linked to the cluster leads to higher test positivity rates. The R_t_ downtrended 30 March onwards, with the most recent estimated R_t_ of 1·159 (95% CI 1·128–1·189) on 22 April which was the lowest value of R_t_ yet. Since there was no significant susceptible depletion, this decrease in transmission can be attributed to the intensive social restrictions in place. The trend of R_t_ from 23 April onwards is of particular interest and it remains to be seen, as to whether the R_t_ can reach sub-threshold levels (below 1) before the lockdown is relaxed.

India is under one of the strictest lockdowns in the world for more than five weeks now,^52^ and a comprehensive lockdown exit strategy will be needed to consolidate and build upon the gains achieved this far. A sudden and complete lifting of the ongoing nationwide lockdown is not a feasible option since it will lead to a rapid exponential increase in cases due to absence of herd immunity. A lockdown of adequate length and efficacy eventually causes the active cases to peak and then gradually decrease. Once the peak of active cases is reached, extending lockdown farther beyond the peak may have additional benefits due to progressive exhaustion of the infectious pool in the population, which is practically comparable to a lower pre-relaxation prevalence of COVID-19. This has 2 effects– first, the rebound epidemic growth is initially slower which delays the resurgent rise in cases after relaxation of lockdown. This seems to imply that though extending lockdown inherently buys time for preparation, it also adds progressively longer preparation time after the lockdown is relaxed (Figure 4A,B). Second, we find that if lockdown is to be reimposed after a fixed relaxation period, the magnitude of the second peak can be reduced by relaxing the lockdown farther from the first peak (Figure 4C,D). This is of particular interest if an intermittent lockdown strategy is implemented in the future, where measures need to be imposed and relaxed repeatedly. The time gained should be used to strengthen surveillance systems, ramp-up testing capacity and increase health-system preparedness. It is optimal to prevent a second wave from occurring at all, by fine-tuning lockdown relaxation based on serial monitoring of R_t_ to keep its value under 1.^6,8^ In this scenario, a later relaxation will allow stabilisation of disease prevalence at a lower value, which can provide a buffer for response if and when a resurgent rise in cases is seen (maintaining R_t_=1 implies that the prevalence will remain constant at the pre-relaxation level). These observations may increase the benefit of lockdowns above what is widely known and can better inform the delicate balance of cost and benefits of such intensive policies.

Massive scaling up of testing has been proposed as a lockdown exit strategy.^17,53^ In this study, we present quantitative evidence based on modelling for the same (Figure 5). Extremely low transmission rates during intensive restrictions are inherently enough to contain the epidemic. However, as transmission rates increase with progressive restoration of normal socio-economic activities post lockdown relaxation, testing assumes an increasingly substantial role in containment. The extent of relaxation that will be possible without causing an untenable rebound in infections, will highly depend on the amount of testing that is done, especially after lockdown relaxation. While having both intensive social distancing policies and very expansive testing may be nonviable, combining the effects of both to a feasible extent can effectively keep the epidemic under control (Figure 5D). Our findings align with results seen in countries with an aggressive testing approach, like South Korea and Taiwan where severe restrictions have been avoided. ^54^ Even if the amount of testing being done during lockdown is deemed to be sufficient, a rapid and massive scaling up of testing capacity is needed preferably before relaxing restrictions. The monetary cost of expanding testing even at a large scale, is expected to be smaller than the cost of implementing intensive social distancing for long periods.^11^ In addition to supporting the economy, this approach can ameliorate the humongous social and humanitarian implications of imposing population-wide lockdowns, especially in a country such as India. ^12,13^

Blanket testing of HCWs can be a judicious use of the expanded capacity, considering they are highly exposed personnel, and risk spreading the infection to patients, co-workers and family members if infected. This will limit depletion of an already scarce workforce due to unnecessary quarantine, while also reducing spread from unrecognised asymptomatic infections in HCWs.^55^ Other essential workforce like law enforcement personnel, grocery vendors, sanitation workers, etc with high contact rates should also be considered.

SARS-CoV-1 did not reach the scale of SARS-CoV-2 despite a comparable R_0_ due to low community transmissibility and onset of infectivity well after symptom onset which allowed optimal efficacy of traditional control measures like symptom-triggered isolation and contact tracing.^19,47^ Pre-symptomatic transmission occurs before the onset of symptoms in an eventually symptomatic patient, while asymptomatic transmission occurs through patients who never become symptomatic. Presence of both these features in COVID-19 is a significant deterrent for control strategies.^16,19,37,56^ In such a scenario and R_0_~2.5, modeling studies indicate that controlling COVID-19 outbreaks through classical contact tracing and isolation alone is not possible.^15^ However, contact tracing systems should be strengthened since they are a prerequisite for expanded testing of contacts, and they may achieve significant containment at lower effective reproduction numbers.^15^ Contact definitions should include contacts made 48-72 hours before symptom onset of index case to account for pre-symptomatic transmission. Technology enabled contact tracing can reduce delay to isolation of contacts thus cutting off transmission when infectiousness is highest around the time of symptom onset.

Based on our findings, it is possible that detecting more asymptomatics through testing impedes transmission to an extent where the total number of infections and thus the number of symptomatic cases decreases (Figure 5), relieving burden upon the healthcare system and reducing mortality. This finding will increasingly approximate reality if asymptomatics play a larger role in transmission. A case in point is a blanket testing study done in a small town of Italy which achieved almost complete outbreak control.^17^ Although blanket testing is not practical for larger implementation, it further highlights the importance of detecting and isolating asymptomatics in controlling COVID-19 outbreaks.

A symptom-based monitoring approach during quarantine will miss asymptomatic infections, who will escape the quarantine net and go on to spread the disease. With emerging evidence of infectious asymptomatics, it is prudent to modify the public health response to address these concerns. Thus, all contacts should ideally be tested at the end of quarantine irrespective of symptoms. In settings where testing all contacts is not yet possible, close contacts may be tested, and extended quarantine periods upto 28 days may be considered, which have two-fold benefits. First, almost all asymptomatics finish their infectious period before 28 days, and second, more symptomatics can be detected by day 28 (only 2 out of 10000 symptomatic cases are missed by day 28, compared to 101 cases by day 14).^29^ Such extended quarantines are already in place in certain parts of India and China.^57,58^ Currently, a 14 day quarantine is recommended based on studies of incubation period of COVID-19,^29,42^ but studying the incubation period inherently assumes an onset of symptoms. It is encouraging to note that the need for expanded testing can be supported by high-throughput machines and by pooling of samples.^41^ Pooling can also be used for community surveillance and has the potential to drastically increase detection capabilities while saving costs and resources. Pooling should be used wherever possible, while also enhancing research to boost pool size and accuracy.^59^

While contact tracing, isolation and testing are important, the role of behaviour change in reducing transmission must not be underestimated. Asymptomatic people are themselves less likely to take appropriate precautions, and people use less caution around other people who don’t have symptoms. Universal mask wearing in public spaces should be encouraged, and mandated by policy if required.^60^ Since ensuring long-term compliance of citizens to health advisories and public restrictions will be another challenge, transparent and proactive communication by authorities along with continued social support for vulnerable groups will be essential.

There is looming uncertainty regarding the burden of asymptomatics and the role they play in transmission. Estimates for asymptomatics range from 18% to 80% of total infections.^17,18,32–34^ With India and China reporting asymptomatics in the higher ranges, it may be possible that young and developing countries have a high proportion of asymptomatic carriers. These estimates are cross-sectional, and thus do not differentiate asymptomatics from pre-symptomatics. It is only retrospectively that the true burden of asymptomatics can be ascertained through serological studies, which will also help us to understand the true fatality rate of COVID-19.^61^ Epidemiologic and virologic studies have established that asymptomatics are infectious, and have similar upper respiratory viral loads as symptomatic patients.^16,17,30,31^ This is in comparison to influenza, where asymptomatics have lower viral loads and thus are less infectious.^19^ Studies to evaluate pre-symptomatic transmission of COVID-19 have shown that 25–50% of total transmission occurred before the index case showed symptoms,^31,56^ although quantitative evidence of asymptomatic transmission is lacking and deserves further research. There is an urgent need to identify these gaps in the understanding of SARS-CoV-2 in order to grasp the true size and severity of this pandemic and plan future strategies accordingly.

Blanket interventions have been effective to suppress the pandemic till now, but targeted interventions will be key as we move forward. Various interventions need to be stratified based on how effectively they suppress viral transmission and the amount of disruption they cause. Cost effectiveness analysis must be done, and bundles of interventions that together achieve high efficacy with least accompanying disruption should be deployed. Highly effective and disruptive interventions should be targeted to areas with active hotspots and high community transmission. It will be essential to build robust disease surveillance systems to assess the relative impact of each intervention in real-time and reduce the time delay to response. Expanded testing and strengthened contact tracing will enable this by reducing the reporting lag and rapidly detecting any surge in cases. Instead of adopting an intermittent lockdown policy, where lockdowns are treated as either ‘on’ or ‘off’,^9^ some countries have adopted a staged alert system for responding to the COVID-19 pandemic,^62^ where a geographical area may move up and down alert levels, to reflect the level of suppression that the local outbreak situation demands. Similarly, India has recently stratified its districts into red, orange and green zones based on surveillance trends, in preparation for a staggered relaxation of lockdown.^63^ Such social distancing policies which are dynamic with respect to geography and time are direly needed as we move into a time of relative uncertainty post lockdown relaxation.

Several limitations of our study should be noted. First, the estimation of reproductive numbers is based upon detected cases which are only a fraction of the actual infections, and we do not account for variation in detection which if significant, may confound the changes in estimated R_t_ over time. Second, we assumed the delay from symptom onset to confirmation to be similar to the delay from symptom onset to hospitalisation due to lack of data, and that this delay is uniform across India. Though the latter approximates the former, further studies to ascertain the true reporting lag in India are needed since it is critical for identifying R_t_ changes at correct points in time.^6^ Third, our model does not factor in pre-symptomatic transmission, which along with a short estimated reporting lag may underestimate the transmission by symptomatic cases and thus inflate the effect of detecting asymptomatics. Fourth, since our primary goal was to evaluate the effect of identifying asymptomatics, we assumed that increased testing increases the detection of asymptomatics only, while in reality it would detect more cases across the clinical spectrum. However, the interpretations regarding impact of increased testing are not sensitive to this assumption, which has been discussed through an alternate interpretation of the model (appendix p4). Fifth, we assumed a constant death rate (κ), in contrast to reality where the death rate gradually decreases during an epidemic to ultimately converge at the near-actual death rate ^61^. Thus, we refrained from forecasting deaths due to obvious bias in the prevailing death rate at the time of the study.

Notwithstanding the limitations, this study estimates reporting lag for COVID-19 in India for the first time, which can be used in future modelling studies. Here, we built a mathematical model which can account for the dynamics of lockdown imposition and relaxation, varying levels of case detection, lag to symptom onset and case reporting, while simultaneously allowing to test the range of asymptomatic burden and transmissibility. Since we have presented findings across the range of uncertainty regarding asymptomatics (appendix), our results are robust to emerging evidence. Though our model is fitted to data from India, we expect the insights into lockdown relaxation and testing impact to be generalizable to similar scenarios elsewhere.

In conclusion, though disruptive, the world’s largest lockdown in India has been effective in reducing the transmission levels of COVID-19. To avoid a resurgence in cases, a dynamic relaxation approach guided by regional monitoring of effective reproduction numbers is recommended, and this relaxation should be farther from the peak of active cases as feasible. Asymptomatic infectives could be a considerable challenge to long-term containment efforts, and increased detection will play an increasingly pivotal role once restrictions start to be lifted. The amount of testing will dictate the extent of resumption of socio-economic activities, and authorities should scale-up testing capacity as a priority. Further, control measures should be appropriate in the social context of a population, as this pandemic brings a humanitarian crisis in addition to a public health one, especially in vulnerable populations across the world.

## Data Availability

The code for the simulations and detailed results are available at the following GitHub repository: https://github.com/technosap/SEIR_QDPA-COVID_19. The primary data for calculation of reporting lag is available in the appendix. All other data is from publicly available datasets.

## Contributors

MG and GGP conceptualised the study. MG and SSM designed the model. SSM programmed the model and performed formal analyses. MG wrote the original draft. SSM, AR, MAg, MAr and AMa assisted in manuscript preparation and reviewed existing evidence. GGP, AL, PB and AB consulted on the analyses and reviewed the draft. SSM and MG made the figures. RK, VPM, PT, SB and AMo collected and curated primary patient data and enabled inter-department coordination at the hospital. GGP and MG supervised the project. All authors interpreted the results, contributed to writing the manuscript, and approved the final version for submission.

## Acknowledgements

We thank all members of the India COVID-19 Apex Research Team (iCART) for sharing their expertise. We thank our professors at the All India Institute of Medical Sciences (AIIMS), New Delhi for their mentorship. SSM received a KVPY fellowship and support from IISER Pune. We thank Dr Jitender Kumar Meena from AIIMS New Delhi and Dr Hemant Deepak Shewade from the International Union Against Tuberculosis and Lung Disease (The Union) for their valuable inputs. The views expressed in this publication are those of the authors and not necessarily those of their affiliated institutes. We express our gratitude to all personnel who are at the frontlines of this pandemic across the globe.

## Ethics Approval

All relevant ethical guidelines have been followed; necessary IRB and/or ethics committee approvals have been obtained (through *Institute Ethics Committee AIIMS Delhi; Ref. No. IECPG-166/23.04.2020)*. All necessary patient/participant consent has been obtained.

